# Host Determinants of Adjuvant Chemotherapy Initiation and Survival in Upper Gastrointestinal Adenocarcinoma

**DOI:** 10.64898/2026.10.13.26365477

**Authors:** Belinda Jiao, Sami Alotaibi, Khalid Abumelha, James Tankel, Lorenzo Ferri, Jamil Asselah, Thierry Alcindor, Carmen L. Mueller, Mathieu Rousseau, Sara V. Soldera, Lawrence Lee, Nicholas Bertos, Mehrnoush Dehghani, Kim Anh Ma, Jonathan Cools-Lartigue

**Author notes:** Corresponding author: Jonathan Cools-Lartigue, MD, PhD, Division of Thoracic and Upper GI Surgery, McGill University Health Centre 1650 Cedar Ave., Room L8.505, Montreal, QC H3G 1A4, Canada, |. Kim Anh Ma and Jonathan Cools-Lartigue contributed equally as senior authors.

## Abstract

**Background:** Successive advances in perioperative systemic therapy have improved outcomes in gastroesophageal adenocarcinoma, yet a substantial proportion of patients fail to complete treatment, and omission of postoperative therapy has been associated with poorer survival. We assessed whether perioperative changes in CBC-derived neutrophil-to-lymphocyte ratio (cNLR) and albumin added information about treatment initiation beyond tumour pathology and whether initiation was associated with survival.

**Methods:** This retrospective cohort included 243 patients undergoing resection at the McGill University Health Centre from 2006 to 2025. Sequential logistic models evaluated pathological characteristics, pretreatment host variables and perioperative changes in cNLR and albumin. Postoperative measurements preceded adjuvant initiation. Nested comparisons used matched complete cases. Day-90 landmark and time-varying Cox models assessed associations with overall survival (OS) and disease-free survival (DFS).

**Results:** Seventy-eight patients (32.1%) did not initiate adjuvant therapy. Adding host variables improved discrimination beyond pathology (AUC 0.672 to 0.763; P=.002). Greater perioperative cNLR increase was associated with lower odds of initiation (adjusted OR 0.67 per unit increase in Δlog₂ cNLR, 95% CI 0.47–0.95), whereas more favourable albumin change was associated with higher odds (OR 1.07 per g/L, 95% CI 1.01–1.13). Neither marker remained independently associated when modelled together. Initiation by day 90 was associated with longer OS (HR 0.53, 95% CI 0.36–0.78) and DFS (HR 0.57, 95% CI 0.39–0.84).

**Conclusions:** Host inflammatory and albumin changes added information about adjuvant initiation beyond tumour pathology, and initiation was associated with longer survival. Prospective evaluation should determine whether host-directed perioperative strategies improve postoperative treatment delivery.

## INTRODUCTION

Perioperative systemic therapy improves outcomes in locally advanced gastroesophageal adenocarcinoma. MAGIC and FNCLCC/FFCD established a survival advantage over surgery alone; FLOT4 improved survival with fluorouracil, leucovorin, oxaliplatin and docetaxel; and MATTERHORN improved event-free survival by adding durvalumab to FLOT^1–4^. These advances emphasize the importance of understanding patients’ ability to receive treatment across the perioperative course.

Delivery of the postoperative component remains challenging. Surgical morbidity, impaired recovery and treatment intolerance may prevent patients from initiating or completing planned therapy. Observational studies associate postoperative chemotherapy after neoadjuvant treatment with improved survival in selected patients, although its contribution is difficult to separate from patient selection ^5,6^. Surgical complications may further limit treatment receipt ^7^. Understanding which patients can resume systemic therapy therefore requires consideration of both residual tumour burden and the patient’s postoperative condition. Identifying host factors associated with treatment delivery could help inform supportive strategies aimed at improving recovery and access to postoperative treatment.

Routine CBC and serum albumin measurements offer accessible, repeatable measures through which to examine this host state. Complete blood count-derived neutrophil-to-lymphocyte ratio (cNLR) captures the relative abundance of circulating neutrophils and lymphocytes and has been associated with prognosis in gastric cancer, including after adjustment for tumour stage ^8^. Albumin provides information about the systemic response to illness: its concentration is influenced by inflammation, altered hepatic protein synthesis and vascular permeability, and low levels are associated with adverse outcomes. It should therefore be interpreted within the clinical context rather than as a direct measure of nutritional status. Serial assessment of these markers may help characterize aspects of recovery that are not captured by tumour pathology or performance status alone ^9^.

Measurement timing is central to this interpretation. Pretreatment values characterize the host before cancer-directed therapy, whereas preoperative values may reflect neoadjuvant treatment and postoperative values capture the subsequent course following resection. Previous studies have associated treatment-related changes and persistent postoperative elevation of NLR with adverse outcomes in gastroesophageal cancers ^10–13^. Whether perioperative cNLR and albumin changes provide additional information about adjuvant treatment initiation beyond pathological and baseline clinical characteristics remains less clear.

We therefore examined tumour and host factors associated with adjuvant systemic therapy initiation in a retrospective cohort of patients undergoing resection for gastroesophageal adenocarcinoma. We evaluated whether perioperative changes in cNLR and albumin added information beyond pathological and pretreatment characteristics and assessed the association between adjuvant initiation and survival. Exploratory analyses examined factors associated with postoperative marker levels, including whether sustained preoperative marker abnormalities provided information beyond the last preoperative measurement.

## METHODS

### Study design and population

We conducted a retrospective cohort study of adults with gastroesophageal adenocarcinoma undergoing curative-intent resection at the McGill University Health Centre (MUHC), Montreal, Canada, from January 1, 2006, to September 30, 2025. Eligibility required histologically confirmed adenocarcinoma and sufficient data to establish treatment delivery. Patients with metastatic disease, early-stage disease treated with surgery alone, or no resection were excluded; nonmetastatic gastroesophageal junction tumours staged IVA, including N3M0 disease, remained eligible. The MUHC Research Ethics Board approved the study (2022-7947) and waived informed consent given the retrospective, deidentified design. All eligible patients meeting the study criteria during the study period were included; no a priori sample-size calculation was performed.

### Data collection and outcomes

Demographic, clinical, pathological and treatment data were abstracted from institutional records and linked to longitudinal CBC and biochemistry data. Race and ethnicity were not systematically documented; country/region of origin was abstracted from charts and harmonized into geographic categories. Recorded variables included regimens, cycle counts, and dates of diagnosis, treatment, surgery, recurrence, death and last follow-up.

The primary outcome was adjuvant systemic therapy initiation (≥1 postoperative cycle). Secondary outcomes were OS (diagnosis to death) and DFS (surgery to recurrence or death); patients without an event were censored at last follow-up, and those with insufficient follow-up were excluded from DFS. Treatment exposure and documented reasons for non-initiation were summarized descriptively; regimen-specific cycle targets for DCF and FLOT were evaluated exploratorily.

### Longitudinal assessment of systemic inflammation

cNLR was the absolute neutrophil count divided by absolute lymphocyte count from the same CBC. Pretreatment baseline cNLR and albumin were the last eligible values within 30 days before first cancer-directed treatment (neoadjuvant initiation, or surgery date for upfront surgery) and strictly before surgery. Preoperative reference values were the last eligible cNLR within surgery days −90 to −1 and albumin within days −30 to −1; postoperative values were the eligible measurement closest to day 30 (days 7–60). Perioperative change was log₂(postoperative/preoperative cNLR) or postoperative minus preoperative albumin (g/L); patients whose postoperative measurement fell on or after adjuvant initiation were excluded from dynamic analyses without reselection.

Preoperative threshold occupancy (cNLR ≥4, based on the published median cutoff^14^; albumin <35 g/L) was the proportion of supported observation time above/below threshold, using linear interpolation, over a primary window of days −30 to −1 (sensitivity: −90 to −1). Full laboratory selection, interpolation and coverage rules, and measurement-window definitions, are in Table S1 and the Supplementary Methods; data availability is reported in Tables S2–S3.

### Adjuvant initiation and survival analyses

Factors associated with adjuvant initiation were evaluated using sequential logistic regression models (Model A: pathology; Model B: + age, ECOG, baseline albumin and cNLR; Models C1– C3: + perioperative Δlog₂ cNLR and/or albumin change; full specifications in Table S4). Model A used standard maximum-likelihood logistic regression; Models B–C3 and their matched comparators used Firth-penalized regression. Nested models were compared on identical complete cases using penalized likelihood-ratio tests and apparent AUCs.

OS and DFS were estimated by Kaplan–Meier methods and compared by log-rank test; median follow-up used reverse Kaplan–Meier analysis. Pathological-stage comparisons were restricted to neoadjuvant-treated, M0, stage-assignable patients; TRG was evaluated across its four categories and as 0–1 versus 2–3.

The principal treatment–survival analysis used a postoperative day-90 landmark: patients alive and under observation beyond the landmark were included (DFS additionally required recurrence-free M0 disease), with adjuvant exposure defined by initiation on or before day 90. Cox models, beginning follow-up at the landmark, adjusted for pathological T category, nodal status, grade, age, clinical albumin and ECOG status; five-year estimates were conditional on landmark eligibility.

A separate exploratory analysis assessed seven factors (pathological stage, TRG, age, and baseline/postoperative cNLR and albumin) using univariable and multivariable Firth-penalized logistic regression in the same complete-case population, with odds ratios per 10-year age increase, cNLR doubling, and 5 g/L albumin increase (Figure 2C). Exploratory analyses also compared adjuvant regimens at day 90 and cycle-target attainment at day 180; full specifications and sensitivity analyses appear in Table S1 and the Supplementary Methods.

**Figure 1.**
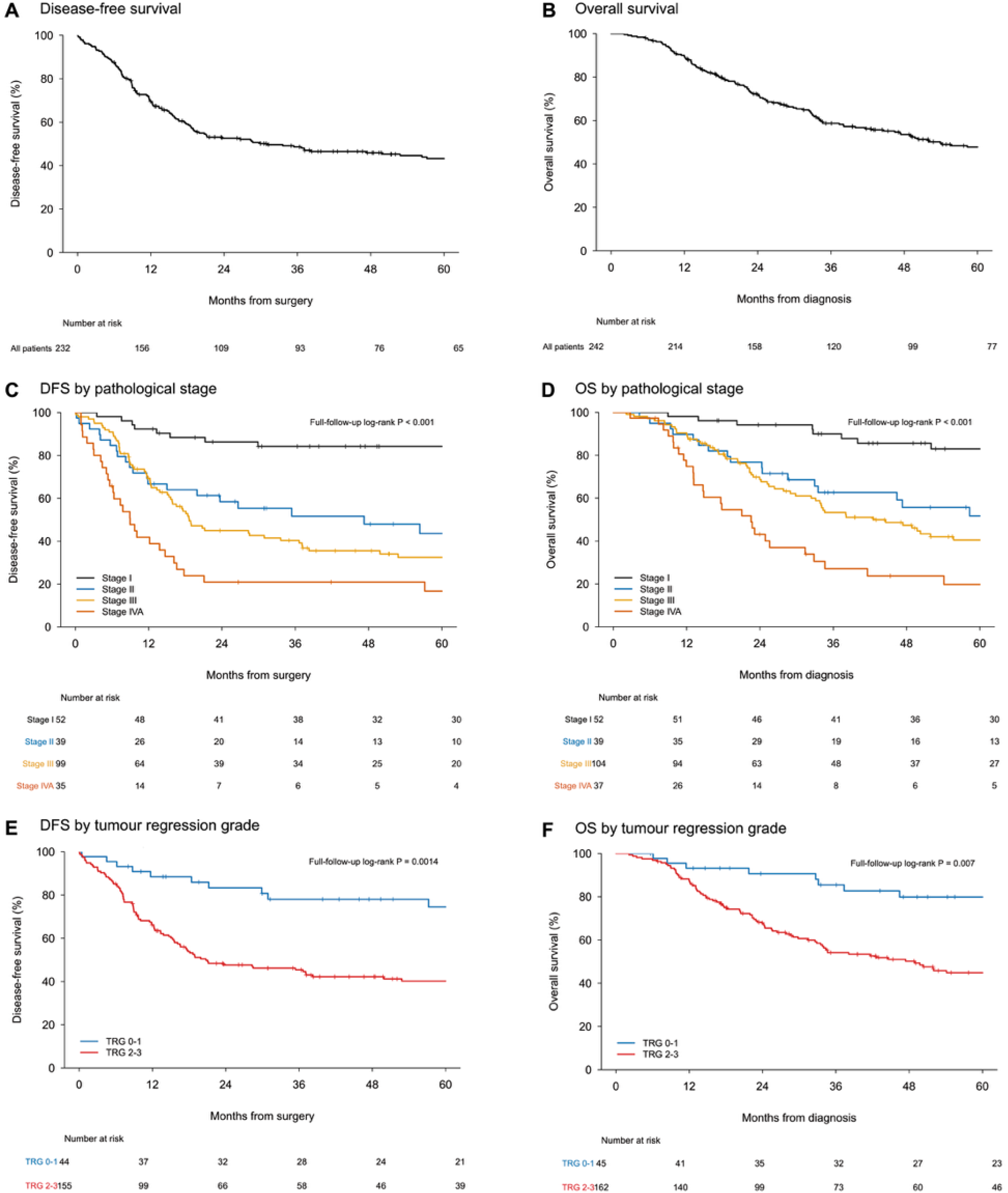
Survival according to pathological stage and tumour regression grade. Kaplan– Meier estimates of disease-free survival (DFS) and overall survival (OS) in the evaluable cohort (A, DFS, N = 232; B, OS, N = 242), by pathological stage (C, DFS, N = 225; D, OS, N = 232), and by tumour regression grade (TRG; E, DFS, N = 199; F, OS, N = 207). DFS was measured from surgery to first recurrence or death, and OS from diagnosis to death from any cause. Stage analyses included neoadjuvant-treated patients with M0 disease and an assignable pathological stage; stage IVA represents nonmetastatic oesophageal/gastroesophageal junction disease. TRG analyses compared scores 0–1 (blue) with 2–3 (red) among neoadjuvant-treated patients with an available score. Curves are displayed through five years; Kaplan–Meier estimation and log-rank comparisons use all available follow-up. Tick marks indicate censoring; tables show numbers at risk.

**Figure 2.**
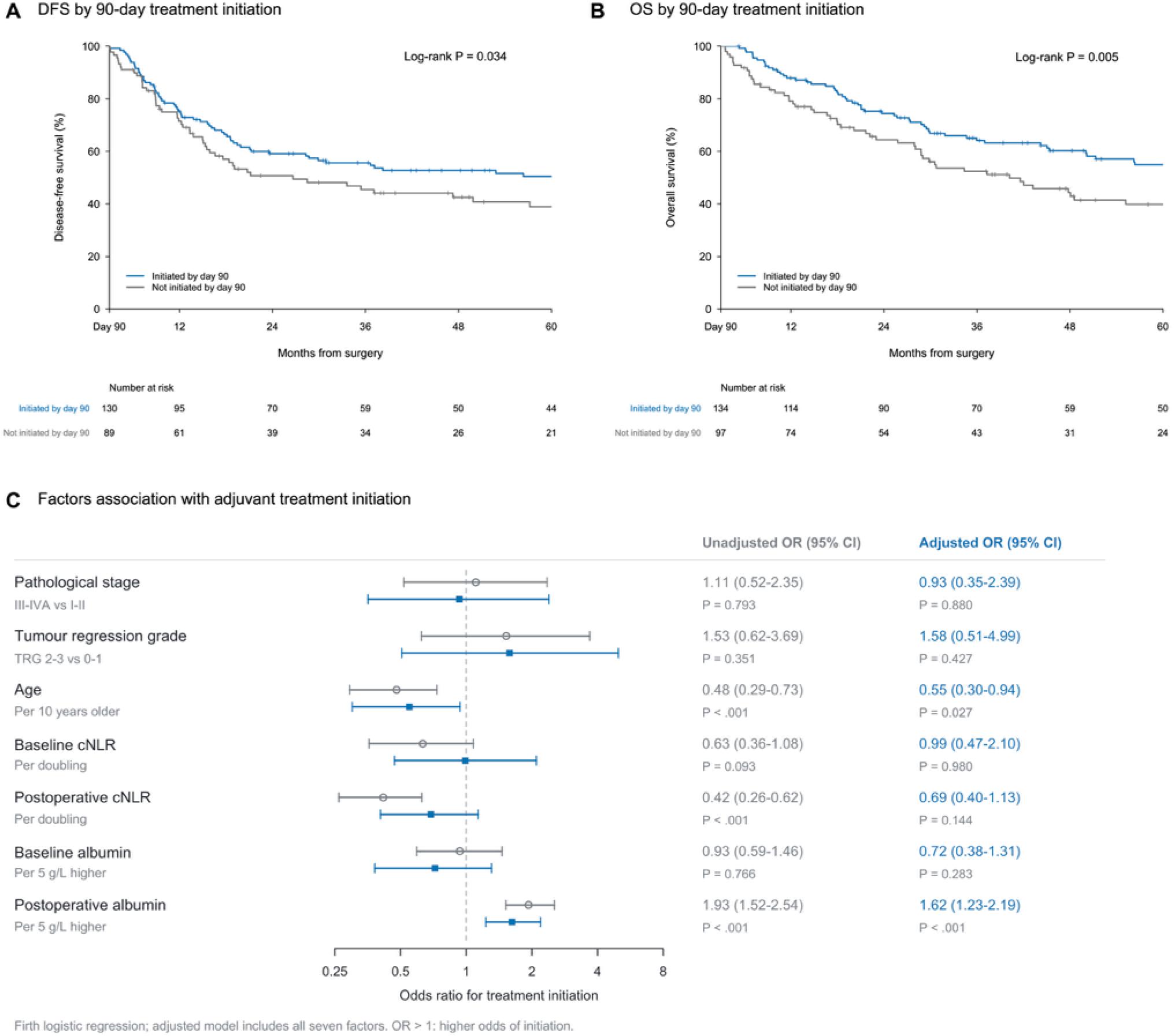
Adjuvant treatment initiation, survival and associated factors. (A–B) Kaplan–Meier estimates of disease-free survival (DFS; A; N = 219) and overall survival (OS; B; N = 231) according to adjuvant initiation by postoperative day 90. Blue indicates initiation by day 90; grey indicates no initiation by day 90, including subsequent initiators. Group sizes were 130 versus 89 for DFS and 134 versus 97 for OS. Patients were alive and observed beyond the landmark; DFS additionally required M0 disease and freedom from recurrence. Curves are displayed through five years after surgery; estimation and log-rank tests use full follow-up. Tick marks indicate censoring; tables show numbers at risk. (C) Unadjusted (grey circles) and mutually adjusted (blue squares) odds ratios for any adjuvant initiation in 119 complete cases, estimated using Firth-penalized logistic regression. The adjusted model includes all seven displayed factors. Bars indicate 95% profile-likelihood confidence intervals. Baseline measurements were selected within treatment days −30 to 0, strictly before surgery; postoperative measurements were selected closest to day 30 within days 7–60 and required to precede initiation. OR >1 indicates higher odds of initiation. cNLR, CBC-derived neutrophil-to-lymphocyte ratio; TRG, tumour regression grade.

### Statistical analysis

Continuous variables were summarized as medians (IQR) and categorical variables as counts (%); analyses used complete cases without imputation, and all tests were two-sided with 95% CIs.

Postoperative log₂ cNLR and albumin were modelled separately by linear regression, adjusting for the corresponding preoperative marker, sampling day, age, ECOG, preoperative BMI, tumour site, pathological T category and nodal status (secondary models added grade, TRG, perineural invasion, length of stay or threshold occupancy), using HC3-robust estimates. Benjamini– Hochberg correction covered the 12 primary clinical-factor comparisons and Holm correction the four occupancy comparisons; other comparisons were exploratory with nominal P values.

Analyses used Python (NumPy, pandas, SciPy, scikit-learn); full diagnostics and sensitivity analyses are in the Supplementary Methods.

## RESULTS

### Cohort characteristics and treatment patterns

The cohort comprised 243 patients who underwent surgical resection (Figure S1). Median age was 64 years (IQR, 58–71), and 206 patients (84.8%) were male. ECOG performance status was 0 or 1 in 235 patients (96.7%). Median recorded albumin was 39 g/L (IQR, 36–42), baseline cNLR was 3.3 (IQR, 2.5–4.4) and preoperative BMI was 26.6 kg/m² (IQR, 24.0–30.4). Median perioperative weight loss was 7.2 kg (IQR, 4.2–11.0; Table 1).

**Table 1.** Patient characteristics (N = 243)

| Characteristic | Value | Available n |
| --- | --- | --- |
| Age, years | 64.0 (58.0–71.0) | 243 |
| Albumin, g/L | 39.0 (36.0–42.0) | 242 |
| Baseline cNLR | 3.3 (2.5–4.4) | 219 |
| Preoperative BMI, kg/m<sup>2</sup> | 26.6 (24.0–30.4) | 240 |
| Postoperative BMI, kg/m<sup>2</sup> | 23.9 (21.1–27.2) | 228 |
| Perioperative weight loss, kg | 7.2 (4.2–11.0) | 228 |
| Male | 206 (84.8%) | 243 |
| Female | 37 (15.2%) | 243 |
| ECOG 0 | 135 (55.6%) | 239 |
| ECOG 1 | 100 (41.2%) | 239 |
| ECOG ≥2 | 4 (1.6%) | 239 |
| ECOG missing | 4 (1.6%) | — |
| **Country/region of origin, n (%)** |  | **243** |
| Canada | 180 (74.1%) | — |
| Europe | 22 (9.1%) | — |
| Other/unspecified | 14 (5.8%) | — |
| Unknown | 9 (3.7%) | — |
| Middle East/West Asia | 4 (1.6%) | — |
| East Asia | 4 (1.6%) | — |
| Latin America/Caribbean | 4 (1.6%) | — |
| South Asia | 2 (0.8%) | — |
| Africa | 2 (0.8%) | — |
| United States | 2 (0.8%) | — |

Values are median (interquartile range) or n (%). Categorical percentages use n=243; available n denotes nonmissing observations. Country/region of origin came from chart abstraction and is not self-reported race or ethnicity. BMI was calculated from recorded height and weight. Perioperative weight loss was preoperative minus postoperative weight; measurement intervals were not standardized. Albumin sampling was not uniformly timed. Baseline cNLR used the last eligible CBC within 30 days before first cancer-directed treatment and strictly before surgery; the latest same-day CBC was selected. ANC and ALC had to be positive; 43 patients contributed treatment-day CBCs. ECOG, Eastern Cooperative Oncology Group performance status; BMI, body mass index; cNLR, CBC-derived neutrophil-to-lymphocyte ratio; ANC, absolute neutrophil count; ALC, absolute lymphocyte count.

For analysis, tumours were grouped as oesophageal/gastroesophageal junction in 186 patients (76.5%) and gastric, including Siewert III tumours, in 57 (23.5%). Most patients had cT3 disease at presentation (223, 91.8%). Surgical pathology demonstrated nodal involvement in 159 patients (65.4%) and grade 3 histology in 107 (44.0%). Tumour regression grade (TRG) was evaluable in 207 neoadjuvant-treated patients: 45 (21.7%) had TRG 0–1 and 162 (78.3%) had TRG 2–3. TRG was not recorded in 31 patients who received neoadjuvant treatment and was not applicable in five patients who underwent upfront surgery. Tumour characteristics are detailed in Table 2.

**Table 2.** Tumour characteristics and pathological stage (N=243)

| **Characteristic** | **N = 243**<br>**n (%)** |
| --- | --- |
| **Tumour location** |  |
| Oesophageal/GEJ | 186 (76.5) |
| Gastric, including Siewert III | 57 (23.5) |
| **Composite pathological stage<sup>1</sup>** |  |
| I | 53 (21.8) |
| II | 41 (16.9) |
| III | 105 (43.2) |
| IVA | 41 (16.8) |
| Gastric complete response, no formal stage group | 3 (1.2) |
| **Pathological T category** |  |
| (y)pT0 | 23 (9.5) |
| (y)pT1–2 | 68 (28.0) |
| (y)pT3–4 | 151 (62.1) |
| (y)pT2/T3, indeterminate between groups | 1 (0.4) |
| **Pathological N category** |  |
| (y)pN0 | 84 (34.6) |
| (y)pN1 | 56 (23.0) |
| (y)pN2 | 47 (19.3) |
| (y)pN3 | 56 (23.0) |
| **Histological grade** |  |
| Grade 1–2 | 110 (45.3) |
| Grade 3 | 107 (44.0) |
| Unavailable | 26 (10.7) |
| **Tumour regression grade<sup>2</sup>** |  |
| TRG 0 | 23 (9.5) |
| TRG 1 | 22 (9.1) |
| TRG 2–3 | 162 (66.7) |
| Not recorded after neoadjuvant treatment | 31 (12.8) |
| Not applicable: no neoadjuvant treatment | 5 (2.1) |

Values are n (%), with all 243 patients as the denominator.
<sup>1</sup> Site-specific AJCC eighth-edition stage groups are summarized descriptively: Siewert I–II follow the oesophageal/GEJ schema and Siewert III the gastric schema. Substages I–III are combined. The prefix yp applies after neoadjuvant treatment (n = 238), and p after upfront surgery (n = 5). Stage IVA is nonmetastatic oesophageal/GEJ disease; metastatic stage IV is separate. Three gastric ypT0N0M0 complete responses have no formal assigned stage group. Gastric N3a/b are combined as N3.
<sup>2</sup> TRG not recorded (n = 31) is distinct from not applicable without neoadjuvant treatment (n = 5). Unrecorded TRGs occurred after resections in 2006–2010 (n = 19) and 2011–2017 (n = 12).

Neoadjuvant systemic therapy was administered to 238 patients (97.9%), whereas 165 (67.9%) initiated adjuvant systemic therapy and 78 (32.1%) did not. Among the 78 patients who did not initiate adjuvant therapy, the primary documented reason was host- or recovery-related for 41 (52.6%), disease-related for 18 (23.1%), other for eight (10.3%), and unknown or undocumented for 11 (14.1%; Table S5). Both treatment phases were received by 162 patients (66.7%). DCF and FLOT were the most frequently administered regimens, accounting for 95 (39.9%) and 71 (29.8%) neoadjuvant treatments, respectively, and 61 (37.0%) and 39 (23.6%) adjuvant treatments. DCF combined with immunotherapy was administered to 41 neoadjuvant recipients (17.2%) and 23 adjuvant recipients (13.9%). Regimen distributions and recorded treatment exposure are summarized in Table 3.

**Table 3.** Systemic treatment delivery (N = 243)

| Measure | Neoadjuvant | Adjuvant |
| --- | --- | --- |
| **Treatment initiated** | 238 (97.9) | 165 (67.9) |
| **Treatment not initiated** | 5 (2.1) | 78 (32.1) |
| **Regimen among recipients** |  |  |
| FLOT | 71 (29.8) | 39 (23.6) |
| FLOT plus durvalumab | 1 (0.4) | 0 (0.0) |
| FOLFOX | 3 (1.3) | 9 (5.5) |
| DCF | 95 (39.9) | 61 (37.0) |
| DCF plus immunotherapy | 41 (17.2) | 23 (13.9) |
| Carboplatin/paclitaxel | 12 (5.0) | 2 (1.2) |
| Docetaxel + cisplatin | 6 (2.5) | 9 (5.5) |
| Cisplatin + 5-FU | 5 (2.1) | 6 (3.6) |
| Other | 4 (1.7) | 16 (9.7) |
| **Recorded cycles, median (IQR)** | 3.0 (3.0–4.0); n = 238 | 3.0 (3.0–4.0); n = 162 |

Values are n (%) unless otherwise specified. Percentages for treatment initiation use all 243 patients; regimen percentages use patients who initiated neoadjuvant (n = 238) or adjuvant (n = 165) treatment. Both treatments were initiated by 162 patients (66.7%).
Other neoadjuvant regimens (n = 4): cisplatin + 5-FU ± pembrolizumab (trial assignment uncertain); cisplatin + 5-FU + trastuzumab ± pertuzumab; DCF followed by cisplatin + 5-FU + trastuzumab; and EOX (epirubicin, oxaliplatin and capecitabine), one patient each.
Other adjuvant regimens (n = 16): CAPOX (capecitabine and oxaliplatin), n = 3; docetaxel + 5-FU, n = 3; docetaxel alone, n = 2; 5-FU + leucovorin, n = 2; 5-FU + leucovorin with radiotherapy, n = 3; capecitabine with radiotherapy, docetaxel + cisplatin + trastuzumab, and FOLFOX + nivolumab, one patient each.
5-FU, fluorouracil; FLOT, fluorouracil, leucovorin, oxaliplatin and docetaxel; FOLFOX, fluorouracil, leucovorin and oxaliplatin; DCF, docetaxel, cisplatin and fluorouracil; IQR, interquartile range.

### Survival outcomes according to pathology and treatment delivery

Median follow-up was 85.0 months. Among 242 patients evaluable for OS, 136 deaths occurred; median OS was 53.9 months (95% CI, 40.0–77.3), and five-year OS was 47.8%. Among 232 patients evaluable for DFS, 140 recurrence or death events occurred; median DFS was 30.9 months (95% CI, 19.0–57.2), and five-year DFS was 43.3% (Figures 1A-B).

Pathological stage was associated with OS and DFS in the post-neoadjuvant M0 cohort (n=232 and n=225, respectively; both global log-rank P<.001). Five-year OS declined from 83.0% for stage I to 51.7%, 40.5% and 19.8% for stages II, III and IVA; corresponding DFS estimates were 84.2%, 43.6%, 32.4% and 16.7% (Figure 1C–D). Stage IVA comprised nonmetastatic oesophageal/gastroesophageal junction tumours. TRG was also associated with OS (n=207; four-group global P=.046) and DFS (n=199; four-group global P=.013). When dichotomized, patients with TRG 0–1 had higher five-year OS and DFS than those with TRG 2–3 (79.9% versus 44.9%, log-rank P=.007; and 74.5% versus 40.2%, P=.0014, respectively; Figure 1E–F).

Adjuvant therapy began a median of 70 days after surgery (IQR, 57–85). At the day-90 landmark, 134 of 231 OS-evaluable patients had initiated treatment, compared with 97 who had not initiated by day 90. Among 219 patients evaluable for DFS, 130 had initiated by day 90 and 89 had not. Five-year OS, conditional on day-90 landmark eligibility, was 54.5% versus 40.6%, and five-year DFS was 50.0% versus 39.7%. Using all available follow-up, initiation by day 90 was associated with longer OS (log-rank P=.005) and DFS (P=.034; Figure 2A–B). After adjustment for pathological T category, nodal status, grade, age, recorded clinical albumin, and ECOG performance status, initiation by day 90 remained associated with longer OS (HR 0.53, 95% CI 0.36–0.78; P=.001; n=202) and DFS (HR 0.57, 95% CI 0.39–0.84; P=.005; n=190).

Treating initiation as a time-varying exposure yielded similar associations with OS (adjusted HR 0.46, 95% CI 0.30–0.69; P<.001) and DFS (HR 0.61, 95% CI 0.40–0.93; P=.021). The OS association persisted with additional stratification by tumour site, nodal status and grade, whereas the DFS association was attenuated.

In exploratory adjusted day-90 regimen analyses, FLOT versus DCF estimates were inconclusive for OS (HR 0.83, 95% CI 0.38–1.81) and DFS (HR 0.93, 95% CI 0.42–2.08). DCF plus immunotherapy had a lower estimated DFS hazard than DCF (HR 0.28, 95% CI 0.09–0.84), whereas the OS estimate remained imprecise.

Among adjuvant DCF and FLOT recipients, 49 of 61 (80.3%) and 28 of 39 (71.8%), respectively, received at least the regimen-specific target number of cycles. In postoperative day-180 landmark analyses adjusted for regimen, pathological T category, nodal status, age and surgery year, with stratification by tumour site, reaching the cycle target was not clearly associated with OS (HR 0.56, 95% CI 0.27–1.13; P=.106; n=95) or DFS (HR 0.71, 95% CI 0.33–1.53; P=.385; n=87).

Restricting OS analysis to patients who remained recurrence-free at the landmark yielded a similar estimate (HR 0.54, 95% CI 0.24–1.18).

### Factors associated with adjuvant treatment initiation

Sequential models evaluated pathological and host factors associated with postoperative chemotherapy initiation (Table 4). The dataset included 12,958 CBC records from 243 patients; 12,886 supported cNLR calculation. Laboratory availability is summarized in Tables S2 and S3.

**Table 4.** Pathology and host factors associated with postoperative chemotherapy initiation

| Model | Specification | Analysed N | Initiated / did not | Apparent AUC | Matched comparator AUC<sup>1</sup> | Incremental P<sup>1</sup> |
| --- | --- | --- | --- | --- | --- | --- |
| **A** | pT, node positivity, grade, TRG and PNI | 179 | 122 / 57 | 0.654 | — | — |
| **B** | A + age, ECOG, baseline albumin and baseline cNLR | 146 | 94 / 52 | 0.763 | A: 0.672 | .0023 |
| **C1** | B + $\Delta\log_2$ cNLR | 126 | 80 / 46 | 0.818 | B: 0.799 | .020 |
| **C2** | B + Δalbumin | 76 | 50 / 26 | 0.810 | B: 0.770 | .011 |
| **C3** | B + $\Delta\log_2$ cNLR + Δalbumin | 75 | 49 / 26 | 0.828 | B: 0.765 | .016<sup>2</sup> |

The pathology-only model (Model A), comprising pT, nodal status, grade, TRG and PNI, included 179 patients and had an apparent AUC of 0.654. Adding age, ECOG, baseline albumin and baseline cNLR (Model B) increased the apparent AUC from 0.672 to 0.763 among the same 146 complete cases (likelihood-ratio P=.0023).

Perioperative changes in cNLR and albumin each provided additional information when added separately to Model B. In Model C1, comprising 126 patients, adding Δlog₂ cNLR increased the apparent AUC from 0.799 to 0.818 (incremental P=.020). A greater postoperative increase in cNLR was associated with lower odds of chemotherapy initiation (adjusted OR 0.67 per unit increase in Δlog₂ cNLR, 95% CI 0.47–0.95; P=.026; Table 5). In Model C2, comprising 76 patients, adding postoperative-minus-preoperative albumin change increased the apparent AUC from 0.770 to 0.810 (incremental P=0.011). A smaller albumin decline or greater increase was associated with higher odds of initiation (adjusted OR 1.07 per 1 g/L higher change, 95% CI 1.01– 1.13; P=0.020).

**Table 5.** Adjusted associations of the dynamic markers

| Model | Dynamic marker | Adjusted OR (95% CI) | Wald P |
| --- | --- | --- | --- |
| **C1** | $\Delta\log_2$ cNLR, per unit<sup>3</sup> | **0.67 (0.47–0.95)** | .026 |
| **C2** | Δalbumin, per 1 g/L | **1.07 (1.01–1.13)** | .020 |
| **C3** | $\Delta\log_2$ cNLR, per unit<sup>3</sup> | 0.69 (0.41–1.17) | .167 |
| **C3** | Δalbumin, per 1 g/L | 1.04 (0.97–1.11) | .273 |

<sup>1</sup> Comparator models were refitted on the same patients as the corresponding expanded model. Incremental P values are penalized likelihood-ratio tests. AUCs across different sample sizes should not be directly compared. Model A's standalone estimate uses ordinary logistic regression; B–C3 and their matched comparisons use Firth regression.
<sup>2</sup> Joint test of both dynamic terms, 2 degrees of freedom.
<sup>3</sup> A one-unit increase represents a doubling of the postoperative/preoperative cNLR ratio. Baseline LDH is excluded throughout B–C3. Baseline albumin and cNLR are retained. $\Delta$ albumin is postoperative minus preoperative albumin. Postoperative values are closest to day 30 within days 7–60 and strictly precede adjuvant initiation; preoperative windows are –30 to –1 days for albumin and –90 to –1 days for cNLR. ECOG is constant and therefore omitted in C2/C3 and their matched comparators. All AUCs are apparent; analyses are exploratory.

The combined dynamic model (C3) included 75 patients. Joint addition of both markers increased the apparent AUC from 0.765 to 0.828 compared with Model B fitted to the same patients (joint incremental P=0.016). However, neither dynamic term showed a clear independent association when included together: the adjusted OR was 0.69 for Δlog₂ cNLR (95% CI 0.41–1.17; P=0.167) and 1.04 for albumin change (95% CI 0.97–1.11; P=0.273). These exploratory estimates reflect apparent, in-sample discrimination; incremental comparisons used matched patient subsets.

In a separate exploratory analysis, seven pathological and host factors were examined simultaneously for their associations with adjuvant treatment initiation (Figure 2C). Unadjusted and mutually adjusted models were estimated in the same 119 complete cases, of whom 76 initiated adjuvant treatment and 43 did not. Older age was associated with lower odds of initiation (adjusted OR 0.55 per decade, 95% CI 0.30–0.94; P=.027), whereas higher postoperative albumin was associated with greater odds of initiation (adjusted OR 1.62 per 5 g/L, 95% CI 1.23– 2.19; P<.001). Higher postoperative cNLR was associated with lower odds of initiation in unadjusted analysis (OR 0.42 per doubling, 95% CI 0.26–0.62; P<.001), but this association attenuated after mutual adjustment (adjusted OR 0.69, 95% CI 0.40–1.13; P=.144). The remaining factors showed no clear independent associations.

### Exploratory factors associated with postoperative cNLR and albumin

Postoperative cNLR and albumin levels were examined in separate exploratory regression models adjusted for the corresponding preoperative marker value, postoperative sampling day, age, ECOG performance status, preoperative BMI, tumour site, pathological T category and nodal status.

Older age was associated with higher postoperative cNLR and lower albumin. Each additional decade corresponded to 15% higher cNLR (ratio 1.15; 95% CI, 1.05–1.27; P=.005; n=189) and 1.83 g/L lower albumin (95% CI, −3.19 to −0.46; P=.009; n=123), although neither association met the 5% false-discovery threshold. In separate secondary models, longer hospitalization was associated with higher cNLR and lower albumin, while grade 3 histology was associated with higher cNLR. Other evaluated clinical and pathological factors showed no clear associations. Hospitalization was considered a correlate because it could overlap marker assessment.

Higher preoperative albumin was associated with higher postoperative albumin (+0.40 g/L per preoperative g/L; 95% CI, 0.11–0.69; P=.008), whereas the last preoperative cNLR showed no clear independent association with postoperative cNLR (ratio per doubling, 1.08; 95% CI, 0.97– 1.21; P=.140).

In the broader 90-day preoperative occupancy analysis, each additional 25 percentage points of covered time with cNLR ≥4 was associated with 13% higher postoperative cNLR after adjustment for the last preoperative value and the other covariates (ratio 1.13; 95% CI, 1.04–1.23; P=.006; n=179). This association remained significant after correction across four marker-window comparisons (Holm-adjusted P=.026). The primary 30-day occupancy analysis was directionally similar but inconclusive (ratio 1.16; 95% CI, 0.99–1.35; P=.070; n=71). Preoperative time with albumin <35 g/L did not demonstrate additional information beyond the last preoperative albumin in either window.

## DISCUSSION

Pathological stage, tumour regression and age remained important clinical determinants. Observational studies similarly associate postoperative treatment delivery with age and postoperative fitness^5,15^. Yet much of the effort to improve perioperative outcomes has concentrated on changing the drugs. MAGIC increased five-year survival from 23% to 36%, and MATTERHORN improved two-year event-free survival from 58.5% to 67.4% by adding durvalumab to FLOT^1,4^. Although systemic regimens have advanced, intensification does not always improve survival; KEYNOTE-585 improved pathological complete response without establishing an overall survival benefit^16^. Ensuring that patients remain sufficiently fit to receive postoperative treatment therefore remains challenging.

That challenge becomes particularly visible after surgery. Adjuvant initiation was documented in only 42.9% of a 2025 real-world FLOT cohort^15^, and approximately one-third of our cohort did not start postoperative therapy. Regimen comparisons were imprecise, whereas initiation was the clearest treatment-related signal. These findings make recovery an important oncologic consideration: residual pathology and age are fixed, and drug access may constrain the available options. However, functional impairment, inadequate intake and reversible complications may remain amenable to intervention.

Our findings identify perioperative recovery as a clinically relevant point of assessment. In the sequential models, rising cNLR and falling albumin were associated with lower odds of adjuvant initiation beyond pathological and pretreatment characteristics. In the separate mutually adjusted analysis, postoperative albumin remained independently associated with treatment initiation, whereas the association with postoperative cNLR was attenuated. Together, these findings suggest that routine perioperative measurements may help identify patients requiring closer assessment and additional support and warrant prospective validation.

Prehabilitation provides a potential intervention. In esophagogastric cancer, exercise and nutritional prehabilitation improved perioperative functional capacity^17^. The randomized PREHAB trial reduced severe complications after colorectal cancer surgery from 29.7% to 17.1%^18^. Exercise during neoadjuvant chemotherapy was also associated with greater tumour regression in a nonrandomized oesophageal study^19^. After adjuvant chemotherapy for colon cancer, the randomized CHALLENGE trial improved DFS (HR 0.72) and OS (HR 0.63)^20^.

Although these interventions and populations differ, they provide a strong clinical rationale for evaluating physical capacity and structured exercise as active components of cancer care.

Prehabilitation may also affect immunity. In 22 patients with oesophageal adenocarcinoma, prehabilitation during neoadjuvant chemotherapy was associated with greater CD8⁺ tumour infiltration and higher tertiary lymphoid structure cell density^21^. Acute exercise studies in breast cancer also demonstrate rapid mobilization of circulating CD8⁺ T cells and NK cells, although transient blood changes do not establish sustained antitumour activity^22^. Our group’s preliminary MMP-LUNG findings similarly suggest changes over a preoperative programme, with improved physical performance accompanied by increased circulating lymphocytes, lower NLR and exploratory shifts in CD8⁺ T-cell populations^23^. These data support investigation of whether prehabilitation-induced systemic and intratumoural immune changes affect treatment tolerance or response.

This study is limited by its retrospective, single-centre design and a treatment era spanning substantial changes in systemic therapy and perioperative care. Patients who initiated adjuvant therapy were likely to differ in fitness, postoperative complications and disease course from those who did not; landmark and time-varying analyses address treatment timing but cannot eliminate this selection. Incomplete characterization of frailty, complications and supportive interventions also limits interpretation of the host associations. Clinically driven laboratory sampling, complete-case selection and small exploratory subgroups introduce further uncertainty. Most importantly, these data identify associations between host state and treatment delivery; they do not establish that modifying cNLR, albumin or physical fitness would improve adjuvant uptake or survival.

## Conclusion

Improving perioperative outcomes requires attention to both treatment efficacy and the host receiving it. Our findings link postoperative inflammatory and albumin changes to adjuvant treatment delivery, while emerging evidence suggests that exercise and nutritional prehabilitation may also influence immune function. Host optimization therefore warrants prospective evaluation for its potential to improve treatment delivery and alter the biological conditions that shape treatment response and disease control.

## Supporting information

STROBE Checklist

## Data Availability

The dataset is not publicly available due to re-identification risk and restrictions imposed by the MUHC Research Ethics Board. Deidentified data may be made available to qualified researchers upon reasonable written request to the corresponding author, subject to institutional ethics approval and a data sharing agreement.

## Acknowledgments

The authors thank the patients of the McGill University Health Centre whose records contributed to this study, and the clinical teams who provided their care. This study was not preregistered with an analysis plan in an independent or institutional registry.

## Ethics approval

This study was approved by the MUHC Research Ethics Board (project 2022-7947). All procedures conformed to the ethical standards of the 1964 Helsinki Declaration and its later amendments. Informed consent was waived given the retrospective deidentified design.

## Author contributions

BJ: Conceptualization, data curation, formal analysis, methodology, writing – original draft, writing – review & editing. SA: Data curation, writing – review & editing. KA: Data curation, writing – review & editing. JT: Writing – review & editing. LF: Writing – review & editing. JA: Writing – review & editing. TA: Writing – review & editing. CLM: Writing – review & editing. MR: Writing – review & editing. SVS: Writing – review & editing. LL: Formal analysis, writing – review & editing. NB: Writing – review & editing. MD: Writing – review & editing. KAM: Conceptualization, supervision, data curation, formal analysis, methodology, writing – original draft, writing – review & editing. JCL: Conceptualization, supervision, data curation, formal analysis, methodology, writing – review & editing.

## Generative AI

Generative artificial intelligence (GPT-5) was used during manuscript preparation to assist with language editing and organization. All scientific content, statistical analyses, interpretations and references were reviewed and verified by the authors, who take full responsibility for the manuscript.

## Previous presentation

Earlier versions of this work were presented at the 2026 ASCO Gastrointestinal Cancers Symposium, San Francisco, CA, USA, and the 2026 ASCO Breakthrough meeting, Singapore, and received Conquer Cancer Merit Awards at both meetings.

## Synopsis

In 243 patients undergoing resection for gastroesophageal adenocarcinoma, perioperative increases in cNLR and decreases in albumin were associated with lower odds of adjuvant therapy initiation; initiation was associated with longer survival.

