## Supplementary material for "Host Determinants of Adjuvant Chemotherapy Initiation and Survival in Upper Gastrointestinal Adenocarcinoma": STROBE Checklist

### STROBE Statement Checklist for Observational Studies

Jiao et al. | Retrospective cohort study | Annals of Surgical Oncology submission

| Section | Item | STROBE recommendation | Where reported |
|---|---|---|---|
| **TITLE AND ABSTRACT** | | | |
| Title And Abstract | **1(a)** | Indicate the study's design with a commonly used term in the title or the abstract. | p. 3 (Abstract, Methods: cohort design); p. 5 (Methods: retrospective cohort study) |
| Title And Abstract | **1(b)** | Provide in the abstract an informative and balanced summary of what was done and what was found. | p. 3 (structured Background, Methods, Results and Conclusions abstract) |
| **INTRODUCTION** | | | |
| Introduction | **2** | Explain the scientific background and rationale for the investigation being reported. | pp. 4-5 (Introduction) |
| Introduction | **3** | State specific objectives, including any prespecified hypotheses. | pp. 4-5 (final Introduction paragraph) |
| **METHODS** | | | |
| Methods | **4** | Present key elements of study design early in the paper. | p. 5 (Study design and population) |
| Methods | **5** | Describe the setting, locations and relevant dates, including periods of recruitment, exposure, follow-up and data collection. | p. 5 (single-centre MUHC cohort; January 1, 2006 to September 30, 2025); pp. 5-7 (exposure and follow-up definitions) |
| Methods | **6(a)** | Give the eligibility criteria and the sources and methods of participant selection. Describe methods of follow-up. | p. 5 (eligibility, exclusions and records); pp. 5-6 (follow-up and censoring); Supplementary Figure S1, p. 4 |
| Methods | **6(b)** | For matched studies, give matching criteria and numbers of exposed and unexposed. | Not applicable - this was not a matched cohort study. Nested model comparisons used matched complete-case subsets, not participant matching (p. 6). |
| Methods | **7** | Clearly define all outcomes, exposures, predictors, potential confounders and effect modifiers. Give diagnostic criteria, if applicable. | pp. 5-7; Supplementary Table S1, p. 5; Supplementary Table S4, p. 8 |
| Methods | **8** | For each variable of interest, give sources of data and details of methods of assessment or measurement. Describe comparability of assessment methods if there is more than one group. | pp. 5-7; Supplementary Methods, pp. 1-3; Supplementary Tables S1-S4, pp. 5-8 |
| Methods | **9** | Describe any efforts to address potential sources of bias. | pp. 5-7 (timing restrictions, identical complete cases, landmark and time-varying analyses, multiplicity correction); p. 11 (limitations); Supplementary Methods, pp. 1-3 |
| Methods | **10** | Explain how the study size was arrived at. | p. 5 (all eligible patients included; no a priori sample-size calculation); Supplementary Figure S1, p. 4 |
| Methods | **11** | Explain how quantitative variables were handled in the analyses. If applicable, describe which groupings were chosen and why. | pp. 5-7; Supplementary Methods, pp. 1-3; Supplementary Tables S1 and S4, pp. 5 and 8 |
| Methods | **12(a)** | Describe all statistical methods, including those used to control for confounding. | pp. 6-7; Supplementary Methods, pp. 1-3 |
| Methods | **12(b)** | Describe any methods used to examine subgroups and interactions. | pp. 6-7 and Supplementary Methods, pp. 2-3 (exploratory subgroup and sensitivity analyses); no formal interaction analysis reported |
| Methods | **12(c)** | Explain how missing data were addressed. | p. 6 (complete-case analyses without imputation); Tables 1-5, pp. 17-20; Supplementary Tables S2-S4, pp. 6-8 |
| Methods | **12(d)** | If applicable, explain how loss to follow-up was addressed. | pp. 5-6 (censoring at last follow-up and reverse Kaplan-Meier estimation) |
| Methods | **12(e)** | Describe any sensitivity analyses. | p. 7; Supplementary Methods, p. 3 |

| Section | Item | STROBE recommendation | Where reported |
|---|---|---|---|
| **RESULTS** | | | |
| Results | **13(a)** | Report numbers of individuals at each stage of the study - for example, numbers potentially eligible, examined for eligibility, confirmed eligible, included in the study, completing follow-up and analysed. | pp. 7-10; Supplementary Figure S1, p. 4; analysis-specific sample sizes in Tables 4-5, p. 20 |
| Results | **13(b)** | Give reasons for non-participation at each stage. | Supplementary Figure S1, p. 4 |
| Results | **13(c)** | Consider use of a flow diagram. | Supplementary Figure S1, p. 4 |
| Results | **14(a)** | Give characteristics of study participants and information on exposures and potential confounders. | pp. 7-8; Tables 1-3, pp. 17-19 |
| Results | **14(b)** | Indicate the number of participants with missing data for each variable of interest. | Tables 1-5, pp. 17-20; Supplementary Tables S2-S4, pp. 6-8; analysis-specific n reported throughout Results |
| Results | **14(c)** | Summarise follow-up time - for example, average and total amount. | p. 7 (median follow-up 85.0 months) |
| Results | **15** | Report numbers of outcome events or summary measures over time. | pp. 7-10; Figure legends, p. 16; Table 3, p. 19 |
| Results | **16(a)** | Give unadjusted estimates and, if applicable, confounder-adjusted estimates and their precision. Make clear which confounders were adjusted for and why they were included. | pp. 8-10; Figure 2 legend, p. 16; Tables 4-5, p. 20; Supplementary Table S4, p. 8 |
| Results | **16(b)** | Report category boundaries when continuous variables were categorized. | pp. 5-7; Figure 2 legend, p. 16; Table 2, p. 18; Supplementary Tables S1 and S4, pp. 5 and 8 |
| Results | **16(c)** | If relevant, consider translating estimates of relative risk into absolute risk for a meaningful time period. | pp. 7-8 and Figure legends, p. 16 (five-year OS and DFS estimates); otherwise not applicable to the odds-ratio analyses |
| Results | **17** | Report other analyses done - for example, subgroup and sensitivity analyses. | pp. 8-10; Supplementary Methods, pp. 2-3 |
| **DISCUSSION** | | | |
| Discussion | **18** | Summarise key results with reference to study objectives. | pp. 10-11 |
| Discussion | **19** | Discuss limitations of the study, taking into account sources of potential bias or imprecision. Discuss both direction and magnitude of any potential bias. | p. 11 (limitations paragraph) |
| Discussion | **20** | Give a cautious overall interpretation of results considering objectives, limitations, multiplicity of analyses, results from similar studies and other relevant evidence. | pp. 10-11 |
| Discussion | **21** | Discuss the generalisability (external validity) of the study results. | p. 11 (single-centre design and long treatment era limitations) |
| **OTHER INFORMATION** | | | |
| Other Information | **22** | Give the source of funding and the role of the funders for the present study and, if applicable, for the original study on which the present article is based. | p. 1 (Funding: None; funder role not applicable) |

Page references correspond to the 20-page manuscript and the separately paginated 9-page supplementary material reviewed on September 24, 2026. STROBE = Strengthening the Reporting of Observational Studies in Epidemiology; MUHC = McGill University Health Centre; OS = overall survival; DFS = disease-free survival.
