## Supplementary files for "Host Determinants of Adjuvant Chemotherapy Initiation and Survival in Upper Gastrointestinal Adenocarcinoma"

### SUPPLEMENTARY METHODS

#### S1. Laboratory selection and threshold-occupancy derivation

Treatment and surgery dates from the clinical master were used to assign laboratory measurements to the windows defined in Table S1. Laboratory availability was assessed separately from eligibility for individual analyses. Tables S2 and S3 report individual measurements and patients with available data; these counts do not impose paired-measurement, treatment-timing or landmark requirements. Zero neutrophil counts were retained in the cNLR availability tally when lymphocyte counts were positive, but the perioperative and occupancy analyses required positive neutrophil and lymphocyte counts. No pseudocounts were added.

Albumin analyses included exact positive numeric results with confirmed collection dates. Results reported using inequality comparators, unconfirmed collection dates or unresolved conflicting values at the same collection timestamp were excluded. For baseline and preoperative reference measurements, the last eligible date within the specified window was selected. Postoperative measurements were selected by proximity to day 30 within days 7–60. Equidistant dates were resolved by selecting the earlier date; multiple measurements on the selected date were resolved using the latest collection timestamp. Measurements were not averaged. Selection preceded application of the adjuvant-timing restriction, and an excluded postoperative measurement was not replaced by an earlier sample. The combined dynamic model required both selected postoperative markers to precede initiation.

Threshold occupancy was calculated separately for cNLR ≥4 and albumin <35 g/L, using the latest eligible measurement on each calendar day. Between successive observations within the exposure window, values were assumed to change linearly on the raw measurement scale. For intervals crossing the threshold, the crossing time was calculated from the interpolated trajectory. Only intervals of ≤28 days contributed to the calculation, and no extrapolation was made before the first or after the last observation. Occupancy was the total supported duration meeting the threshold divided by the total duration of eligible intervals. Patients with a single observation or no eligible interval had undefined occupancy rather than zero occupancy. Albumin values remaining exactly at 35 g/L contributed no time below threshold.

Surgery-day and postoperative measurements were excluded from preoperative occupancy calculations. Measurements after death and CBCs on or after the recorded myelodysplastic syndrome marrow-diagnosis date were excluded, as applicable. The primary occupancy window comprised surgery days −30 to −1; days −90 to −1 formed the broader sensitivity window. The longer window could include neoadjuvant treatment and therefore reflected marker variation during both treatment and the interval preceding surgery.

#### S2. Model specifications and estimation

##### Adjuvant initiation

Sequential logistic model specifications and categorical reference groups are provided in Table S4. Age, albumin, $\log_2$ cNLR and perioperative changes were entered as continuous linear terms. Baseline host measurements in Model B were pretreatment values, distinct from the preoperative references used to derive the dynamic terms. Covariates without variation within a complete-case subset were omitted because their effects could not be estimated.

The standalone pathology-only model (Model A) was estimated using ordinary maximum-likelihood logistic regression. Models B–C3 and all comparator models refitted for matched complete-case comparisons were estimated using Firth-penalized logistic regression. For sequential initiation models A–C3, odds ratios were obtained by exponentiating fitted coefficients, with approximate Wald confidence intervals and coefficient tests based on the inverse information matrix. Incremental penalized likelihood-ratio tests compared the full model with a model constraining the added coefficients to zero while retaining the full model's information penalty. Tests involved one degree of freedom for an individual dynamic marker and two for their joint addition.

Reference and expanded models were fitted to identical patients for each comparison. Consequently, AUC differences between models fitted in different patient subsets were not interpreted as incremental improvements. AUCs were calculated from fitted probabilities and represent apparent discrimination without cross-validation or optimism correction.

Survival, regimen and cycle-target analyses

Kaplan–Meier analyses used all available follow-up. Pathological-stage comparisons included neoadjuvant-treated patients with M0 disease and an assignable stage, pooling stage III subdivisions for the combined stage comparison. TRG was examined across categories 0–3 and as 0–1 versus 2–3. Median follow-up was estimated by reverse Kaplan–Meier analysis.

For day-90 treatment analyses, exposure was fixed according to initiation by the landmark. Subsequent initiation did not change the assigned group. In time-varying analyses beginning at surgery, patients contributed unexposed time until adjuvant initiation and exposed time thereafter. Initiation models adjusted for pT category, nodal status, grade, age, recorded clinical albumin and ECOG performance status. Recorded clinical albumin in these survival models was distinct from the verified pretreatment albumin used in the sequential initiation models. Cox models used the Efron method for tied event times.

Exploratory regimen comparisons were restricted to patients who had initiated DCF, FLOT or DCF plus immunotherapy by postoperative day 90 and met endpoint-specific landmark eligibility. The recorded adjuvant regimen category was used; later regimen changes were not modelled. DCF was the reference, with adjustment for age, pT category, nodal status and surgery year and stratification by tumour site.

Cycle-target analyses were restricted to adjuvant DCF and FLOT recipients. Attainment indicated receipt of at least three DCF or four FLOT cycles; it did not establish full-dose delivery of every agent. Eligibility required a documented treatment end on or after initiation and on or before postoperative day 180, together with endpoint-specific landmark eligibility. Invalid treatment-date sequences were excluded from survival analyses. Models adjusted for regimen, pT category, nodal status, age and surgery year, with stratification by tumour site.

Postoperative marker models

Separate ordinary least-squares models evaluated postoperative $\log_2$ cNLR and albumin. Each model included the corresponding preoperative marker, postoperative sampling day, age, ECOG, BMI, tumour site, pT category and nodal status. ECOG was grouped as ≥1 versus 0 in these analyses, differing from the 0–1 versus ≥2 grouping in the initiation models. Age, BMI and sampling day were scaled per 10 years, 5 $kg/m^2$ and 10 days, respectively. Grade, TRG, perineural invasion and $\log_2$ length of stay were added separately in secondary models.

HC3 heteroskedasticity-robust covariance estimates were used, with t-based intervals and tests using residual degrees of freedom. Exponentiating cNLR coefficients to base 2 yielded adjusted postoperative cNLR ratios. Albumin coefficients remained in g/L. Conditional on the preoperative value, the coefficients for other predictors also describe differences in perioperative change.

Occupancy was entered continuously per 25-percentage-point increase. Models with and without occupancy used identical complete cases and retained the corresponding preoperative marker. Sampling-adjusted models additionally included the number of preoperative sampling days and the fraction of the window supported by eligible intervals. No thresholds were optimized against postoperative outcomes.

### S3. Sensitivity analyses and diagnostics

Ordinary maximum-likelihood logistic regression was used to assess the sensitivity of the initiation findings to Firth penalization. Numerical convergence and finite coefficient standard errors were checked.

Treatment–survival sensitivities included time-varying initiation from surgery; stratification by tumour site, nodal status and grade; additional adjustment for surgery year and sex with tumour-site stratification; a day-120 landmark; exclusion of patients initiating after postoperative day 180; and administrative censoring at five years after surgery. The day-90 OS analysis was also restricted to patients with M0 disease and documented freedom from recurrence at the landmark. Cycle-target OS analyses were repeated among patients recurrence-free at day 180.

Proportional-hazards diagnostics for initiation and cycle-target models used score tests for covariate interactions with log(1 + analysis time), including exposure-specific and global tests. No additional proportional-hazards diagnostic was performed for the exploratory regimen comparisons. Numerical checks assessed Cox-model convergence and finite standard errors.

Postoperative marker sensitivities added a quadratic sampling-day term, adjusted for surgery year, restricted postoperative measurements to days 21–45, and included all available paired measurements with adjustment for whether postoperative sampling occurred on or after adjuvant initiation. Leverage and Cook's distance were examined, and each primary model was repeated after excluding the observation with the largest Cook's distance as an influence diagnostic.

Occupancy sensitivities used same-day medians instead of the latest actual daily value, required at least seven covered days in the 30-day window or 28 covered days in the 90-day window, and additionally adjusted for sampling frequency and coverage. Threshold crossings, exact-threshold values, single observations and exclusion of long gaps were checked against analytical expectations.

Benjamini–Hochberg correction covered the 12 primary clinical-factor tests: age, ECOG, BMI, tumour site, pT category and nodal status for each postoperative marker. The preoperative marker and sampling-day terms were adjustment variables outside this family. Holm correction covered the four added-occupancy tests spanning two markers and two windows. These corrections did not encompass all exploratory analyses conducted during manuscript development; secondary and sensitivity comparisons were reported with nominal P values.

SUPPLEMENTARY FIGURES

**Figure S1. Cohort flow.**

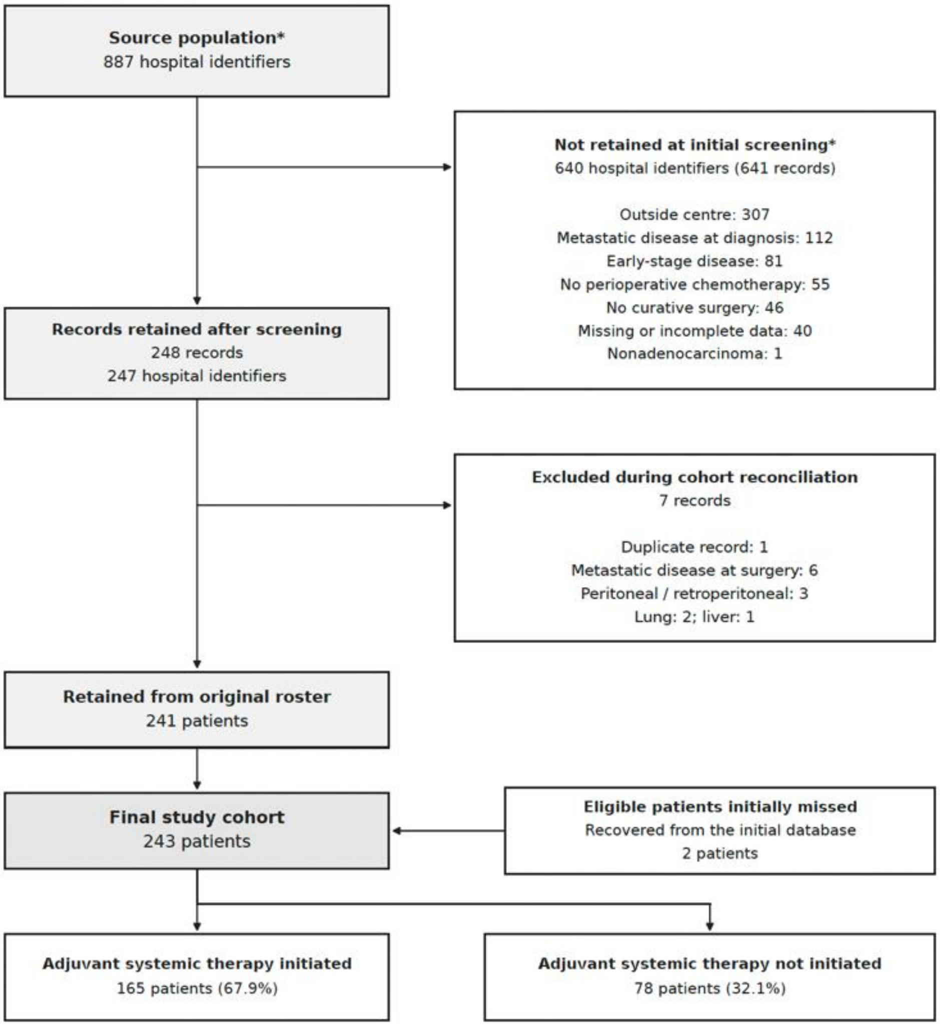


*Exclusion reasons are not mutually exclusive; one excluded record carries two reasons.

SUPPLEMENTARY TABLES

**Table S1. Laboratory exposure windows and analysis eligibility**

| **Measurement or analysis** | **Exposure window or definition** | **Selection and eligibility** |
|---|---|---|
| Pretreatment baseline cNLR and albumin | Days −30 to 0 relative to first cancer-directed treatment | Last eligible measurement within the window, strictly before surgery |
| Preoperative cNLR reference | Surgery days −90 to −1 | Last eligible actual cNLR value |
| Preoperative albumin reference | Surgery days −30 to −1 | Last eligible actual albumin value |
| Postoperative cNLR and albumin | Surgery days 7–60 | Eligible actual value closest to day 30; equidistant dates select the earlier date |
| Perioperative change and postoperative marker analyses | Paired preoperative and postoperative measurements defined above | Selected postoperative measurement must precede adjuvant initiation. Both initiators and non-initiators are eligible; no fixed survival landmark is required |
| Preoperative threshold occupancy | Primary: surgery days −30 to −1; sensitivity: days −90 to −1 | Supported time with cNLR ≥4 or albumin <35 g/L, linked to the eligible postoperative marker. Requires at least two sampling days contributing an eligible interval |
| Adjuvant initiation–survival association | Initiation on or before postoperative day 90 | OS: alive and observed beyond day 90. DFS: additionally recurrence-free. Follow-up begins at day 90; later initiators remain in the not-initiated-by-day-90 group |
| Time-varying initiation–survival analysis | Actual adjuvant initiation date | Follow-up begins at surgery; exposure changes from uninitiated to initiated at the recorded start date |
| Exploratory regimen comparison | DCF, FLOT or DCF plus immunotherapy initiated by postoperative day 90 | Restricted to recipients of these regimens who meet the day-90 OS or DFS eligibility criteria; follow-up begins at day 90 |
| Adjuvant cycle-target analysis | At least three DCF cycles or four FLOT cycles; postoperative day-180 landmark | Requires a valid treatment course documented as ending by day 180, with survival and observation beyond the landmark. DFS additionally requires M0 disease and freedom from recurrence. Follow-up begins at day 180 |

**Timing and measurement rules:** All interval endpoints are inclusive. First cancer-directed treatment is the start of neoadjuvant therapy or, for upfront surgery, the surgery date. Multiple measurements on a selected date are resolved using the latest collection timestamp. Logarithmic cNLR analyses require positive neutrophil and lymphocyte counts from the same CBC. Albumin requires an exact positive result with a confirmed collection date and no unresolved same-time conflict. Postoperative measurements are selected before applying the initiation-timing restriction; excluded measurements are not replaced by an earlier sample.

**Occupancy rules:** Linear interpolation uses the latest actual measurement on each day and only intervals of ≤28 days, without extrapolation beyond observed measurements. Occupancy is the proportion of supported time meeting the threshold. A single measurement does not define occupancy. Sensitivities use daily medians and require at least seven covered days in the 30-day window or 28 covered days in the 90-day window.
Population rules: Complete-case eligibility depends on the covariates required for each analysis. Separate marker analyses do not require availability of the other marker. The day-180 OS cycle-target analysis was also repeated among patients recurrence-free at the landmark.
**Abbreviations:** CBC, complete blood count; cNLR, CBC-derived neutrophil-to-lymphocyte ratio; DCF, docetaxel, cisplatin and fluorouracil; DFS, disease-free survival; FLOT, fluorouracil, leucovorin, oxaliplatin and docetaxel; OS, overall survival.

**Table S2. Availability of albumin, LDH and CRP measurements across treatment and surgical windows**

| Window | Definition | Albumin | LDH | CRP |
|---|---|---|---|---|
| All recorded results | Any date | 241 / 6,346 | 234 / 4,961 | 203 / 1,407 |
| All eligible results | Any confirmed date | 241 / 6,270 | 234 / 4,953 | 203 / 1,381 |
| Baseline | Treatment days −30 to 0; before surgery | 212 / 395 | 146 / 222 | 61 / 75 |
| Pretreatment | On/before first treatment; before surgery | 218 / 657 | 157 / 317 | 87 / 161 |
| Preoperative | Surgery days −30 to −1 | 183 / 287 | 96 / 129 | 29 / 36 |
| Any time before surgery | Surgery days <0 | 227 / 1,761 | 217 / 1,230 | 111 / 276 |
| Postoperative days 0–30 | Surgery days 0–30 | 101 / 233 | 34 / 42 | 70 / 316 |
| Postoperative days 31–90 | Surgery days 31–90 | 190 / 482 | 168 / 375 | 53 / 105 |
| After day 90 | Surgery days >90 | 219 / 3,794 | 204 / 3,306 | 149 / 684 |
| L90 | Surgery days 60–120 | 190 / 570 | 172 / 505 | 41 / 80 |
| Recovery | Surgery days 60–180 | 200 / 906 | 186 / 779 | 64 / 133 |

Eligibility. Window counts include exact positive numeric results with confirmed collection dates and exclude unresolved results with conflicting values at the same collection timestamp. Of 12,714 recorded results, 110 were excluded: 59 with unconfirmed collection dates, 31 reported using < or > comparators, and 20 unresolved conflicting results. The excluded totals were 76 albumin, 8 LDH and 26 CRP records.
Timing. Treatment day 0 is the first neoadjuvant treatment date, or the surgery date when no neoadjuvant therapy was received. Surgery day 0 is the operation date. Baseline and pretreatment results must fall strictly before surgery; preoperative results can follow neoadjuvant treatment. All stated interval endpoints are inclusive.
Counting. Measurements were counted individually without averaging or collapsing to daily values. Patients may contribute to multiple windows. Day 30 is counted only in the 0–30 interval.

L90 and recovery overlap each other and other postoperative intervals; counts should not be summed across overlapping rows.

**Table S3. Availability of CBC measurements and calculable cNLR and SII across treatment and surgical windows**

| Window | Definition | CBC | cNLR | SII |
|---|---|---|---|---|
| All recorded results | Any date | 243 / 12,958 | 243 / 12,886 | 243 / 12,886 |
| All eligible results | Any confirmed date | 243 / 12,958 | 243 / 12,886 | 243 / 12,886 |
| Baseline | Treatment days −30 to 0; before surgery | 219 / 613 | 219 / 612 | 219 / 612 |
| Pretreatment | On/before first treatment; before surgery | 224 / 1,119 | 224 / 1,117 | 224 / 1,117 |
| Preoperative | Surgery days −30 to −1 | 205 / 494 | 205 / 494 | 205 / 494 |
| Any time before surgery | Surgery days <0 | 232 / 2,885 | 232 / 2,883 | 232 / 2,883 |
| Postoperative days 0–30 | Surgery days 0–30 | 231 / 2,955 | 231 / 2,938 | 231 / 2,938 |
| Postoperative days 31–90 | Surgery days 31–90 | 211 / 1,100 | 210 / 1,098 | 210 / 1,098 |
| After day 90 | Surgery days >90 | 229 / 6,018 | 229 / 5,967 | 229 / 5,967 |
| L90 | Surgery days 60–120 | 209 / 1,055 | 209 / 1,053 | 209 / 1,053 |
| Recovery | Surgery days 60–180 | 220 / 1,639 | 220 / 1,637 | 220 / 1,637 |

CBC denotes a recorded complete blood count panel; individual components may be missing. cNLR is the absolute neutrophil count (ANC) divided by the absolute lymphocyte count (ALC). SII, the systemic immune-inflammation index, is the platelet count multiplied by cNLR. ANC, ALC and platelet counts are expressed in $10^9$/L.
cNLR requires a recorded nonnegative ANC and a positive ALC. SII additionally requires a positive platelet count. Of 12,958 CBC records, 72 did not support cNLR calculation: 60 had missing ANC/ALC and 12 had nonpositive ALC. All 12,886 calculable cNLR records also had valid platelet counts, giving identical cNLR and SII availability. Zero ANC values remain eligible for this availability tally; analyses requiring logarithms must separately exclude zero index values.
Sampling days. The CBC master contains 11,919 distinct patient sampling days; calculable cNLR and SII are available on 11,872 patient sampling days. The table reports individual records, without averaging or collapsing multiple measurements on the same day.
Window interpretation. L90 comprises postoperative days 60–120 and recovery comprises days 60–180. Availability in these windows does not itself impose landmark survival, follow-up, recurrence or treatment eligibility criteria. Baseline and pretreatment counts exclude surgery-day samples. Clinical master dates supersede discrepant treatment dates stored in the CBC file.

**Table S4. Sequential logistic regression models for adjuvant systemic therapy initiation**

| Model | Variables included | Purpose |
|---|---|---|
| A: Pathology | Pathological T category, nodal status, histological grade, TRG and PNI | Assess pathological factors associated with initiation |
| B: Baseline host factors | Model A + age, ECOG performance status, baseline albumin and $\log_2$ baseline cNLR | Assess additional information from pretreatment host characteristics |
| C1: Dynamic cNLR | Model B + perioperative $\Delta\log_2$ cNLR | Assess additional information from perioperative cNLR change |
| C2: Dynamic albumin | Model B + perioperative albumin change | Assess additional information from perioperative albumin change |
| C3: Combined dynamic markers | Model B + perioperative $\Delta\log_2$ cNLR and albumin change | Assess the joint contribution of both dynamic markers |

**Definitions:** Pathological variables were coded as pT3–4 versus pT0–2, node positive versus negative, grade 1–2 versus 3, TRG 2–3 versus 0–1, and PNI present versus absent. ECOG was coded as 0–1 versus ≥2. Age and albumin were modelled continuously. Baseline measurements were obtained within days −30 to 0 relative to first cancer-directed treatment and strictly before surgery. $\Delta\log_2$ cNLR = $\log_2$(postoperative/preoperative cNLR); albumin change = postoperative minus preoperative albumin, in g/L. Measurement windows are specified in **Table S1**.

**Analysis:** The outcome was any documented adjuvant systemic therapy initiation. Dynamic models required the selected postoperative measurement to precede initiation; C3 required both postoperative measurements to meet this condition. No fixed day-30 landmark was imposed. Each nested comparison used identical complete cases, with the reference model refitted in that subset. The standalone pathology-only model (Model A) was estimated using ordinary maximum-likelihood logistic regression. Models B–C3 and all comparator models refitted for matched complete-case comparisons were estimated using Firth-penalized logistic regression. Ordinary maximum-likelihood estimates were also examined as sensitivity analyses for the Firth-fitted models. AUCs describe apparent, in-sample discrimination. Covariates without variation within a subset were omitted.

**Abbreviations:** AUC, area under the receiver operating characteristic curve; cNLR, CBC-derived neutrophil-to-lymphocyte ratio; ECOG, Eastern Cooperative Oncology Group; PNI, perineural invasion; TRG, tumour regression grade.

**Table S5. Reasons for adjuvant chemotherapy non-initiation (n = 78)**

| Reason | n (%) |
|---|---|
| **Host- and recovery-related** | |
| Postoperative complications (anastomotic leak, fistula, ICU admission, ARDS) | 24 (30.8) |
| Frailty / poor postoperative recovery | 17 (21.8) |
| **Subtotal** | **41 (52.6)** |
| **Disease-related** | |
| Disease progression or recurrence before initiation | 10 (12.8) |
| Death before initiation | 8 (10.3) |
| **Subtotal** | **18 (23.1)** |
| **Other** | |
| Treatment toxicity | 5 (6.4) |
| Patient refusal | 2 (2.6) |
| Treatment decision / response-guided | 1 (1.3) |
| **Subtotal** | **8 (10.3)** |
| Unknown / not documented | 11 (14.1) |
| **Total** | **78 (100)** |

Values are n (%). Percentages use the 78 patients who did not initiate adjuvant chemotherapy as the denominator and sum to 100.1% because of rounding. ARDS, acute respiratory distress syndrome; ICU, intensive care unit.
